# Regional nonsense constraint offers clinical and biological insights into rare genetic disorders

**DOI:** 10.1101/2024.10.10.24315185

**Authors:** Alexander J. M. Blakes, Nicola Whiffin, Colin A. Johnson, Jamie Ellingford, Siddharth Banka

## Abstract

Understanding the molecular consequences of variants which introduce premature termination codons (PTCs) is essential to predicting their clinical impact^1^. Although transcripts with PTCs are generally expected to undergo nonsense-mediated mRNA decay (NMD)^2–4^, up to 45% of all possible PTCs in the human genome are predicted to escape NMD^5^. Existing studies of constraint against predicted loss-of-function variants at the transcript level^6–10^ do not account for regional differences in NMD efficiency. Here, we developed an NMD-informed constraint metric using sequencing data from 730,947 individuals and found 2,764 transcripts with regional nonsense constraint. In rare disease cohorts, *de novo* nonsense and frameshift variants in constrained regions are up to 9.4-fold enriched and associated with up to 6.4-fold higher odds of diagnosis than in unconstrained regions. We prioritise fourteen candidate disease genes in which constrained regions harbour clusters of nonsense and frameshift variants. These findings will enhance variant interpretation, deliver mechanistic insights, and empower the discovery of novel disease genes.

## Main

To understand the impact of NMD-escaping PTCs on genetic disease (**Figure 1a**) we first annotated predicted NMD-escape regions in the canonical transcripts of all human protein-coding genes. Because the position of the PTC in the mature mRNA is the major determinant of NMD efficiency^11^, we used positional rules to define potential ‘NMD-escape’ regions^12^ (**Figure 1b**). These included ‘start-proximal’ regions^11,13,14^ (<150nt from the canonical start codon), ‘long exon’ regions^11,14^ (>400nt upstream of a splice donor site) and ‘distal’ regions^15^ (the final exon, and the most 3’ 50nt of the penultimate exon). Protein-coding sites outside of these regions were defined as ‘NMD-target’ regions.

**Figure 1.**
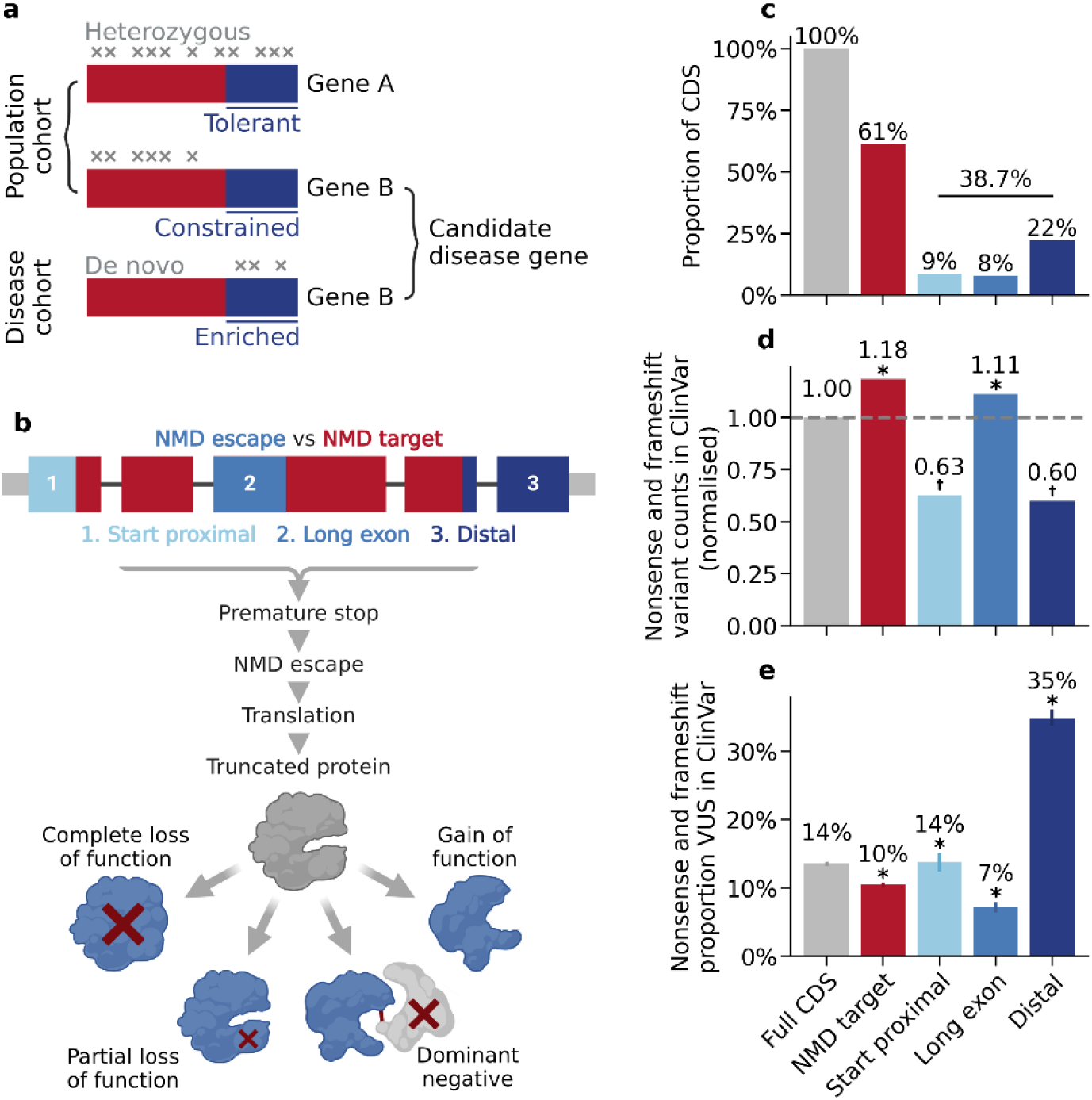
PTCs in predicted NMD escape regions are challenging to interpret clinically. a) Graphical abstract of this study. We identified NMD regions in protein coding genes which are significantly constrained for nonsense variation in population sequencing cohorts. Within these constrained regions, we explored patterns of *de novo* protein truncating variation in rare disease cohorts to prioritise novel candidate disease genes. b) The molecular consequences of NMD escape. Transcript diagram (top) showing coding exons (thick boxes), with NMD-target regions (red) and NMD-escape regions (blue). Introns (black line) and UTRs (narrow grey boxes) are shown (not to scale). Transcripts carrying PTCs in certain regions may escape NMD. The transcript is translated into a truncated protein product. Protein truncation may have diverse consequences. c) The genomic footprint of NMD regions in the canonical transcripts of 19,676 human protein coding genes (Methods). The proportion of the CDS occupied by predicted NMD-escape regions is shown. d) Ascertainment of nonsense and frameshift variants in ClinVar. The absolute variant count in each region is normalised by the genomic footprint of that region. Asterisks indicate categories which are significantly different across all pairwise comparisons (Chi-squared goodness-of-fit test with Bonferroni correction for six tests at alpha=0.05, P<0.0083). Dagger symbols indicate categories which are significant for all pairwise comparisons, except other categories with a dagger symbol. e) The proportion of nonsense and frameshift variants classified as variants of uncertain significance (VUS) in ClinVar. Asterisks indicate categories which are significantly different across all pairwise comparisons (two-sided proportions Z test with Bonferroni correction for six tests at alpha=0.05, P<0.0083).

Overall, 38.7% of coding sequence (CDS) positions in canonical transcripts occur in predicted NMD-escape regions (**Figure 1c**), whereas 61.3% occur in predicted NMD-target regions. To gauge the clinical relevance of these annotations, we examined the frequency and classification of nonsense and frameshift variants in ClinVar^16^. Compared to NMD-target regions, nonsense and frameshift variants in start-proximal and distal regions are both significantly under-reported (**Figure 1d**, Chi-squared goodness-of-fit test, Bonferroni P<0.0083) and significantly enriched for variants of uncertain significance (VUS) in ClinVar (**Figure 1e**, two-sided proportions Z test, Bonferroni P<0.0083). These data highlight the challenge of clinically interpreting nonsense and frameshift variants which are predicted to escape NMD.

To quantify the overall strength of selection against subgroups of nonsense variants, we calculated the mutability-adjusted proportion of singletons (MAPS)^6^ using exome sequencing data from 730,947 individuals in gnomAD v4.1^8^ (**Figure 2a**). We found that nonsense variants in all three NMD-escape regions are constrained, but the constraint was significantly weaker for start-proximal (MAPS=0.083) and distal regions (MAPS=0.066) compared to NMD-target regions (MAPS=0.112) (two-sided proportions Z test, Bonferroni P<0.0024). This is consistent with previous observations that NMD-escaping variants are under weaker negative selection than NMD-targeted variants^11^. Conversely, nonsense variants in long exon regions are significantly more highly constrained (MAPS=0.130, two-sided proportions Z test, Bonferroni P<0.0024), which may reflect the large number of coding positions lost by these truncations. These data show that the strength of selection against nonsense variants can markedly differ depending on the position of the variant.

**Figure 2.**
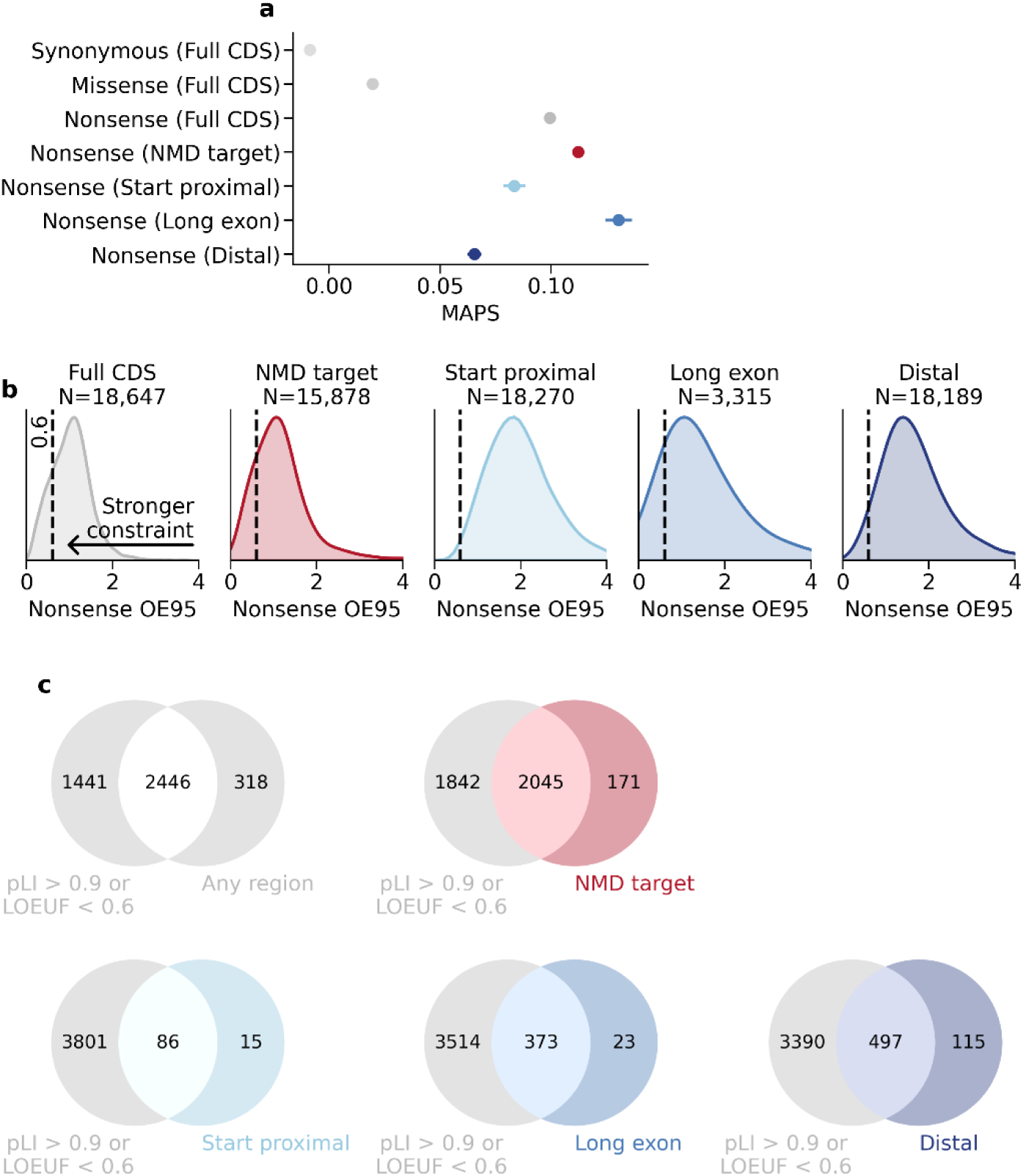
Regional nonsense constraint identifies additional transcripts which are intolerant to PTCs. a) MAPS for nonsense variants in NMD target regions (red) and NMD-escape regions (blue). Error bars show 95% confidence intervals. All categories are significantly different from one another (pairwise two-sided proportions Z tests with Bonferroni correction for 21 tests at alpha=0.05, P<0.0024, asterisks not shown). b) Distributions of the upper bounds of the 95% confidence interval around the observed / expected ratio of nonsense variants (nonsense OE95), stratified by NMD region. The dashed black line marks OE95=0.6. The number of transcripts in which the nonsense OE95 could be quantified is shown. c) Venn diagrams showing the intersection between transcripts with regional nonsense constraint, and transcript-level constraint in gnomAD v4.1 (pLI>0.9 or LOEUF<0.6).

To quantify regional constraint against nonsense variants in individual transcripts, we used mutation rate estimates from Roulette^17^, scaled to the gnomAD v4.1 exome sequencing data, to model the expected number of single nucleotide variants (SNVs) in a given transcript or NMD region. For each transcript, region, and single nucleotide variant consequence (synonymous, missense, or nonsense), we calculated the ratio of observed / expected variants (O/E) (**Extended Data** Figure 1), and the upper 95% confidence interval for this value (OE95) (**Figure 2b**). Transcript-level OE95 values correlate strongly with loss-of-function observed / expected upper fraction (LOEUF) scores^8^ from gnomAD v4.1 (Spearman rho = 0.871, P<10^-8^) thus validating our approach (**Extended Data** Figure 2).

We identified 2,668 / 19,676 (13.6%) canonical transcripts with significant transcript-level nonsense constraint (FDR<0.05, one-sided Poisson-binomial test) (**Extended Data** Figure 3). Applying these models to predicted NMD-target and NMD-escape regions, we found 2,764 / 19,676 (14.0%) transcripts with significant nonsense constraint in at least one region (**Figure 2c**). These included 2,216 (11.3%) transcripts constrained in NMD-target regions, 101 (0.51%) in start-proximal regions, 396 (2.0%) in long exon regions, and 612 (3.1%) in distal regions (FDR<0.05, one-sided Poisson-binomial test) (**Figure 2c**). 507 (2.6%) transcripts are constrained in more than one region (**Extended Data** Figure 3). Importantly, 318/2,764 (11.5%) transcripts with regional nonsense constraint are unconstrained (pLI^6^ <= 0.9 and LOEUF^8^ >= 0.6, N=311) or are missing pLI and LOEUF annotations (N=7) in gnomAD v4.1 (**Figure 2c, Extended Data** Figure 4). These results show that our regional nonsense constraint metric prioritises additional transcripts which are intolerant to PTCs.

To validate our findings, we compared regional nonsense constraint with orthogonal measures of functional importance. Constrained regions are significantly more highly conserved across species (phyloP^18^) and have higher *in silico* predictions of functional importance (AlphaMissense^19^) and pathogenicity (Combined Annotation Dependent Depletion, CADD^20^) than unconstrained regions (**Extended Data** Figure 5, two-sided Welch’s T-tests, Bonferroni P<10^-8^). Surprisingly, relative expression (base-level pext^21^) is comparable between constrained and unconstrained regions (**Extended Data** Figure 5d). Together, these results suggest that constrained regions are more likely to harbour functionally important sites.

Non-uniform distributions of PTCs may explain reduced penetrance of Mendelian disorders in population cohorts^22^ or identify new disease associations or pathomechanisms in known or novel disease genes^23,24^. We therefore examined patterns of nonsense and frameshift variation in transcripts with differential constraint across NMD regions (**Extended Data** Figure 6). We find genes with strong transcript-level nonsense constraint but with NMD regions which are apparently tolerant to PTCs. Examples include *AJAP1* (**Figure 3a**), *GATA6*, and *KANSL1* (**Supplementary** Figure 1-2). We also find genes with weak transcript-level nonsense constraint but strong nonsense constraint in NMD-target regions. Examples include *BTF3* (**Figure 3b**), *FBXL17*, *MNX1*, *PROSER1*, and *RSPO3* (**Supplementary** Figures 3-6). A clinically relevant example is *FOXF1* (Alveolar capillary dysplasia^25^, MIM 265380), which has a clear bimodal distribution of nonsense and frameshift variants in gnomAD, whereas pathogenic nonsense variants in ClinVar have an inverse distribution (**Figure 3c**). Finally, we find genes with weak transcript-level nonsense constraint but strong nonsense constraint in NMD-escape regions, including *CBX7* (**Supplementary** Figure 7), *IRF2BP2* (**Supplementary** Figure 8), and *SLC16A9* (**Supplementary** Figure 9). Genes in the protocadherin alpha cluster (*PCDHA1* to *PCDHA13*) are a striking example; all have weak transcript-level LoF constraint (LOEUF>0.8) but strong nonsense constraint in distal NMD-escape regions (**Figure 3d, Supplementary** Figure 10). The final three exons of *PCDHA1* encode a common cytoplasmic domain shared by all genes in the cluster^26,27^. Distal truncations therefore appear highly deleterious, whereas the non-overlapping long first exons of each gene are tolerant to nonsense and frameshift variants. We observe a similar trend in the protocadherin gamma gene cluster (**Supplementary** Figure 11). Collectively, these results show that regional nonsense constraint captures clinically and biologically relevant patterns of truncating variation.

**Figure 3 (overleaf).**
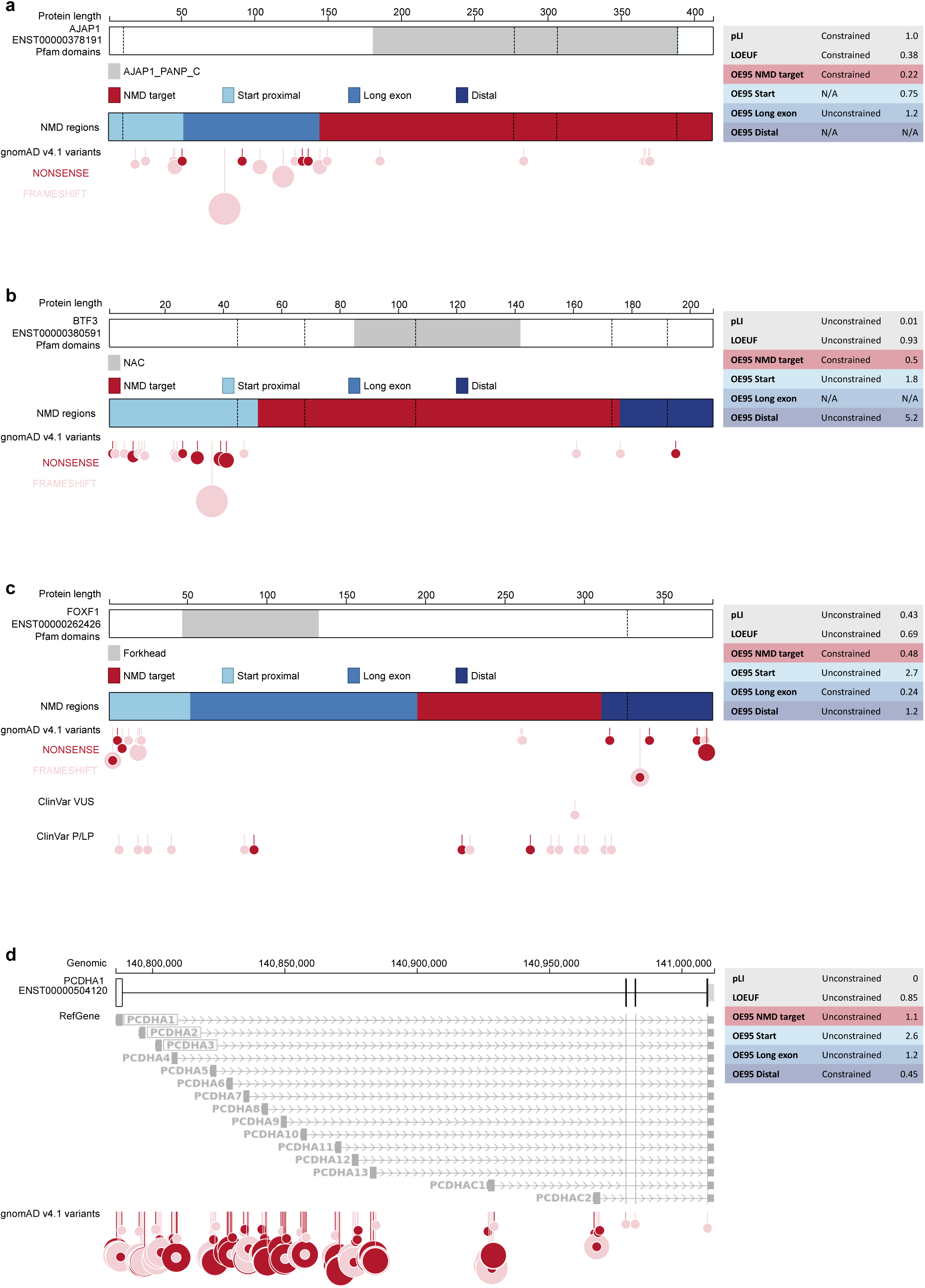
Many transcripts have non-uniform nonsense constraint. **a)** Nonsense variants are not constrained in the long exon NMD-escape region of *AJAP1* (ENST00000378191). The CDS for the MANE Select transcript is shown (thick boxes). Exon junctions are marked by vertical dashed lines. Pfam domains are shown in grey (top); the short name of the domain is given in the legend. NMD- target regions (red) and NMD-escape regions (blue) are shown (middle). Nonsense variants (red circles) and frameshift variants (pink circles) from gnomAD v4.1 are shown (bottom). The size of the circle corresponds to the allele count of the variant. Regional nonsense OE95, pLI and LOEUF scores are shown (right). Note that ENST00000378191 contains a 3’ UTR intron, and therefore the CDS does not contain a distal NMD-escape region. **b)** *BTF3* (ENST00000380591) is constrained for nonsense variants in the NMD-target region. **c)** *FOXF1* (ENST00000262426) nonsense and frameshift variants are bimodally distribution in gnomAD v4.1. Pathogenic nonsense variants in ClinVar are more centrally located. **d)** *PCDHA1* (ENST00000504120) and other protein coding genes in the protocadherin alpha cluster (chr5:140786140-141012347) have strong nonsense constraint in their distal NMD-escape regions. The canonical transcript for every protein coding gene in the cluster is show in grey.

To explore the utility of regional nonsense constraint in clinical variant interpretation, we examined diagnostic outcomes for carriers of *de novo* nonsense or frameshift variants in 13,908 rare disease trios from the 100,000 Genomes Project^28,29^. Because our constraint metric measures selection against heterozygous variants, *de novo* nonsense and frameshift variants in constrained regions are strong *a priori* diagnostic candidates in severe genetic disorders. Individuals carrying a nonsense or frameshift variant in a constrained transcript had significantly greater odds of receiving a diagnosis (OR 4.79, 95% CI 4.04 – 5.67) than those without (**Figure 4a**). Similarly, carriers of *de novo* nonsense or frameshift variants in NMD-target, long exon, or distal regions have significantly higher odds of diagnosis (OR 3.64 (2.97-4.46), 6.42 (4.28-9.63), 5.76 (3.89-8.52) respectively). Importantly, the odds of diagnosis are unchanged for carriers of *de* novo nonsense and frameshift variants in unconstrained regions (**Figure 4a**). Diagnostic outcomes for carriers of *de novo* nonsense and frameshift variants therefore strongly align with regional constraint, highlighting the clinical utility of this metric for variant prioritisation and interpretation in the molecular diagnosis of rare genetic conditions.

**Figure 4.**
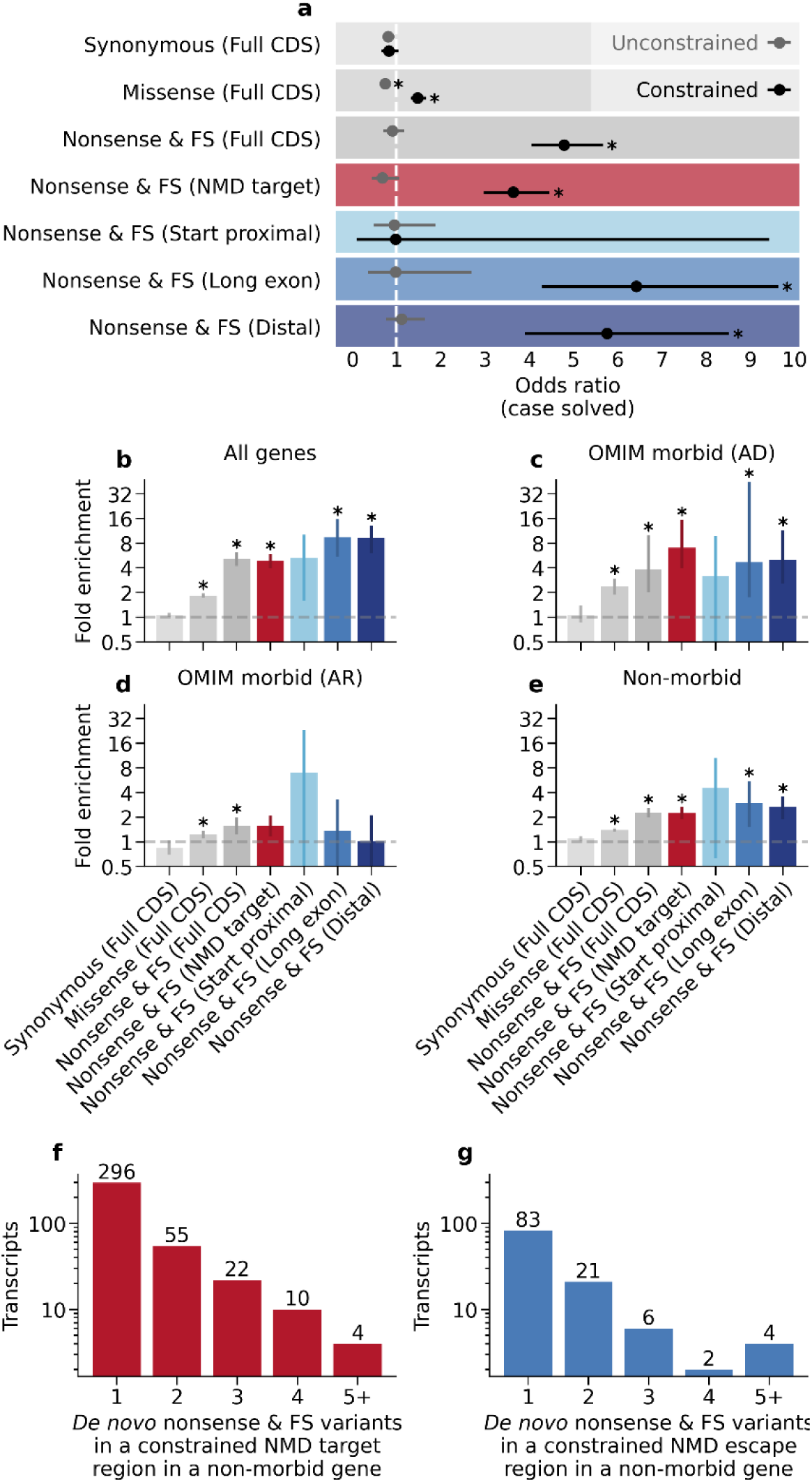
Regional nonsense constraint highlights clinically impactful *de novo* variants and novel disease gene candidates. a) Odds of receiving a diagnosis (“case solved”) for 13,908 rare disease trios in the 100,000 Genomes Project. Error bars represent 95% confidence intervals. Asterisks indicate statistically significant values (two-sided Z test on log transformed values with Bonferroni correction for 14 tests at alpha=0.05, p<0.0036). b) to e) Relative enrichment of *de novo* variants in constrained versus unconstrained regions for 27,762 rare disease trios. Relative enrichment is shown for all genes (b), monogenic disease genes associated with autosomal dominant phenotypes in OMIM (c), monogenic disease genes associated with autosomal recessive phenotypes in OMIM (d) and genes with no monogenic disease association in OMIM (e). Error bars show bootstrap 95% confidence intervals. Asterisks show statistically significant values (percentile bootstrap with Bonferroni correction for 28 tests at alpha=0.05, p<0.0018). f) and g) The number of transcripts with a constrained NMD-target region (f) or NMD-escape region (g) harbouring one or more *de novo* nonsense or frameshift variants in 27,762 rare disease trios. FS = frameshift variant.

Selective constraint can be used to prioritise novel causative genes in monogenic human diseases, including developmental disorders^30,31^. Half of severe developmental disorders are caused by *de novo* variants^32^ and analysis of *de novo* variation in large cohorts has successfully identified novel disease genes and damaging functional variant classes^33,34^. To explore the contribution of PTCs in constrained regions to monogenic disorders, we compared the abundance of *de novo* nonsense and frameshift variants in constrained versus unconstrained regions in large cohorts of rare disease trios (N=27,762 trios). After adjusting for sequence length and mutation rate, we find a substantial enrichment of *de novo* nonsense and frameshift variants in constrained transcripts (5.1-fold enrichment) (**Figure 4b**). Strikingly, this is more pronounced in constrained long exon and distal regions (9.4- and 9.2-fold enrichment respectively) than NMD target regions (4.8-fold enrichment). We subset our analysis by disease annotations in OMIM^35^. *De novo* nonsense and frameshift variants are highly enriched in constrained regions of known dominant disorder genes (**Figure 4c**), but not recessive disorder genes (**Figure 4d**). Importantly, this regional enrichment persists in genes with no disease annotation in OMIM (**Figure 4e**), implying that these variants are an indicator for novel disease genes that remain to be discovered.

To explore novel candidate disease gene associations, we curated genes with clusters of *de novo* nonsense and frameshift variants in constrained regions. In total, we identified 843 genes with at least one *de novo* nonsense or frameshift variant in a constrained region. Of these, we found 48 genes with >=3 such variants in a constrained region, but which lack a dominant monogenic disease association in OMIM (**Figure 4f, 4g, Extended Data Table 1**). We propose that this gene set is likely to include novel monogenic disorder genes. Indeed, several of these genes have recently been associated with developmental disorders (e.g. *FOSL2*^36^, PCBP2^37^, ZFHX3^38^). After excluding genes reported in disease gene curation databases (PanelApp Australia^39^ and Gene2Phenotype^40^) and filtering for regions with nonsense O/E values < 0.3 and fewer than 20 nonsense variants in gnomAD v4.1, we highlight a set of fourteen genes (*ARID4A, BRD1, CLASP1, FBRS*, *HDAC2*, *MGA*, *NAA25*, *NCL*, *RIF1*, *RLF*, *UBR5*, *VEZF1*, *WAPL, ZSWIM8*) as potential candidates for novel disorders. Taken together, these results show that PTC-introducing *de novo* variants in rare disease cohorts are enriched in both NMD-target regions and predicted NMD-escape regions, and that novel disorders driven by heterozygous variants in these regions remain to be discovered.

In conclusion, we apply mutational models to biobank-scale sequencing data to construct a regional nonsense constraint metric offering biological and clinical insights into human genetic disease. We find over 2,000 genes with regional nonsense constraint, show that *de novo* variants and diagnostic outcomes align with constraint in disease cohorts, and prioritise fourteen candidate disease genes. Although our approach is limited to measuring selection against heterozygous variants, does not model indels (a major class of PTC-introducing variants), and depends on prescriptive NMD rules, we suggest that this NMD-aware nonsense constraint annotation may be of considerable utility in studies of PTCs in health and disease.

## Methods

### Representative transcripts

One representative transcript was selected for every protein-coding gene in the GENCODE v39 comprehensive gene annotation^41^. Transcripts were filtered using the following criteria: ‘tag contains “Ensembl_canonical”’, ‘gene_type == “protein_coding”’, ‘transcript_type == “protein_coding”’. 19,676 transcripts meeting these criteria were identified. Where available, the MANE Select transcript (MANE v0.95)^42^ was chosen (N = 18,584). For genes lacking a MANE annotation, the APPRIS principal isoform^43^ was chosen (N = 1,092).

### Defining NMD regions

For each transcript, the following predicted NMD regions were annotated with custom scripts:

- Start-proximal: CDS positions <=150nt downstream of the translation start site.
- Long exon: CDS positions >400nt upstream of an exon-intron junction.
- 50nt rule: CDS positions <=50nt upstream of an exon-intron junction in the penultimate exon (the second-most 3’ exon).
- Last exon: CDS positions in the final exon (the most 3’ exon).
- NMD target: CDS positions lacking any of the above annotations.

The “last exon” and “50nt rule” annotations also account for transcripts which contain 3’ UTR introns, which comprise ∼4% of canonical transcripts. For example, in these transcripts the most 3’ CDS positions do not have a “last exon” annotation. “Last exon” and “50nt rule” annotations were unified into one category: “Distal”. Positions with multiple NMD region annotations were assigned one definitive annotation with this priority: start-proximal > distal > long exon.

### Consequence annotation of coding SNVs

The set of all possible single nucleotide variants (SNVs) in our CDS of interest was annotated with custom scripts using bedtools v2.31.1^44^, BCFtools v1.20^45^, and the GRCh38 reference genome (GenBank assembly accession GCA_000001405.28). SNVs were annotated with the Variant Effect Predictor (VEP) v105^46^ and the Ensembl v105 cache, which is consistent with MANE v0.95^42^ and GENCODE v39^41^. Only autosomal SNVs annotated as “synonymous_variant”, “missense_variant”, or “stop_gained” in the Ensembl canonical transcript for each gene were kept, yielding 22,563,474 synonymous, 71,502,748 missense, and 4,047,794 nonsense SNVs.

### SNVs in gnomAD v4.1

SNVs were identified in exome sequencing data from gnomAD v4.1^47^. Variants were filtered through the gnomAD sample and variant quality control (QC) pipeline described here: https://gnomad.broadinstitute.org/news/2023-11-gnomad-v4-0/. SNVs in our transcripts of interest, which passed all variant filters (FILTER=“PASS”) and had a minimum allele count (AC) of 1, were extracted from the 730,947 samples which passed sample-level QC. In total, 17,885,374 SNVs meeting these criteria were identified. SNVs were labelled with the consequence annotations described above.

### Mutability-adjusted proportion of singletons

For CDS variants observed in gnomAD with median coverage >=20 (N = 17,665,054), we calculated the proportion of singletons (AC == 1) for each consequence (synonymous, missense, nonsense) and NMD region (NMD target, Long exon, Distal, Start proximal) using custom scripts.

Variants in more mutable sequence contexts are more common and have a lower proportion of singletons. To account for sequence mutability, we fit a weighted least squares model of proportion of singletons against scaled mutation rate for synonymous variants in each context. We used mutation rate estimates from Roulette^17^ (see below). There were 99 unique values for the scaled mutation rate among the observed synonymous variants. The model was weighted by the number of possible variants in each variant context.

For each functional variant class, we used the model to predict the expected proportion of singletons. This number was subtracted from the observed proportion of singletons to find MAPS.

For statistical comparison of MAPS between each variant class, all MAPS scores were adjusted by the expected proportion of singletons for synonymous variants before performing pairwise two-sided Z tests of proportion with Bonferroni correction.

### Mutational model for coding SNVs

The expected number of variants among our SNVs of interest was derived from mutation rate estimates from Roulette^17^. Roulette is a base pair level mutation rate model which accounts for local sequence context, DNA methylation, and transcription to accurately model SNV mutation rates genome-wide^17^. Mutation rate estimates from Roulette were scaled to the gnomAD v4.1 exomes to account for differences in cohort size and composition, as described here: https://github.com/vseplyarskiy/Roulette/tree/main/adding_mutation_rate. The set of high quality synonymous sites described in Seplyarskiy et al.^17^ were used as “background” or “neutral” variants for calibrating mutation rate estimates (N=19,969,231). Synonymous variants from the background set were identified among the 17,885,374 gnomAD-observed SNVs described above. Scaled mutation rate estimates were calculated for all possible SNVs genome-wide. We filtered for SNVs in our CDS positions of interest, yielding annotations for 95,668,203 unique SNVs. The scaled mutation rates provide an estimate of the probability of observing each SNV in the gnomAD v4.1 exomes^8^, in the absence of negative selection.

### Regional nonsense constraint

Because transcripts and regions which are poorly covered may appear depleted of nonsense variants (false positives), we only included positions with median sequencing depth >=20 in this analysis.

The proportion of variants expected in each transcript and NMD region was taken as the mean scaled mutation rate for each functional variant class (synonymous, missense, and nonsense variants) in each region. We multiplied this proportion by the number of possible variants, to obtain the number of expected variants in each region.

For each transcript, region, and functional variant class, we treated expected variant counts as a Poisson binomial random variable with the number of trials (N) equal to the number of possible variants, and the probability of observing each variant (P) equal to the scaled mutation rate of that variant under the null model (above).

To identify constrained transcripts and regions, we performed a one-sided test for the number of variants observed against the given Poisson binomial distribution^48^. We tested the null hypothesis that the number of observed nonsense variants is equal to or greater than the number of expected nonsense variants in each transcript and region.

We also calculated the ratio of observed/expected variants (O/E), as well as the upper bound of the 95% confidence interval for the O/E value (OE95). We defined OE95 as the upper bound of the 95% binomial confidence interval for the proportion of variants observed, multiplied by the number of possible variants and divided by the number of expected variants.

After correcting for multiple testing with the Benjamini-Hochberg / false discovery rate (FDR) method^49^, we defined constrained transcripts and regions as those meeting the following criteria:

1. Fewer nonsense variants than expected (one-tailed binomial test, FDR < 0.05)
2. The number of synonymous variants observed is not nominally lower than the number expected (one-sided binomial test, P >= 0.05)
3. The upper bound of the 95% confidence interval of the nonsense O/E value (nonsense OE95) is less than 0.6.

Unconstrained transcripts and regions were defined as follows:

1. Non-significant constraint scores prior to FDR correction (one-tailed Z test, P >= 0.05).
2. At least one nonsense variant observed in the cohort.

Transcripts and regions which did not meet the criteria for “constrained” or “unconstrained” status were annotated as “indeterminate”.

### phyloP, CADD, AlphaMissense, and pext annotations

Base-level phyloP scores from multiple sequence alignments of 241 mammals^18^, Combined Annotation Dependent Depletion (CADD) scores^20^, AlphaMissense scores^19^, and base-level proportion expressed across transcripts (pext) scores^21^ were downloaded from their publicly available repositories. Download URLs are given in the *src/downloads/* directory in the accompanying code repository. CDS sites and variants of interest were annotated with these scores using custom scripts. Site-level AlphaMissense scores were taken as the lowest AlphaMissense score per site per transcript. We successfully annotated 33,834,625 CDS sites with phyloP scores, 93,913,916 CDS SNVs with CADD, 19,900,876 CDS sites with AlphaMissense, and 32,120,976 CDS sites with pext scores.

### Truncating variants in ClinVar

Clinically interpreted variants were obtained from the ClinVar^16^ variant_summary.txt file downloaded on 14/11/23. We filtered variants on the following criteria: Assembly == GRCh38; autosomal variants only; ReviewStatus does not contain either of “no assertion” or “no interpretation”; ClinicalSignificance does not contain any of “not provided”, “drug response”, “other”, “risk”, “low penetrance”, “conflicting”, “affects”, “association”, “protective”, “confers sensitivity”. We identified 2,055,907 unique variants meeting these criteria.

We annotated these variants with VEP v105^46^ and the Ensembl v105 cache. Only autosomal SNVs annotated as “synonymous_variant”, “missense_variant”, “stop_gained”, or “frameshift_variant” in the Ensembl canonical transcript for each gene (where the HGNC^50^ ID of the gene matched the ClinVar annotation) were kept. We successfully annotated 1,575,388 variants. For our analyses of truncating variants in ClinVar we limited these variants to unambiguous “stop_gained” (N=47,216) or “frameshift_variant” (N=79,882) variants.

### Known disease genes in OMIM

A list of known monogenic disease genes was obtained from the OMIM^35^ genemap2.txt file downloaded on 06/11/23. Genes were excluded from the morbid gene list if they had no associated phenotype in OMIM, no mode of inheritance was given for the disorder, no Ensembl gene identifier was available, or if the gene-disease association was annotated as “non-disease”, “susceptibility”, or “provisional”. In total, 3,985 monogenic disease genes were identified.

### *De novo* variants

*De novo* variants (DNVs) overlapping our transcripts of interest were identified in short-read sequencing data from two large cohorts of rare disease trios.

First, we used a set of high-confidence DNVs from whole genome sequencing (WGS) of 13,932 trios in the 100,000 Genomes Project (100KGP)^28^. The annotation pipeline used to identify these variants is publicly available at this URL: https://re-docs.genomicsengland.co.uk/de_novo_data/. We excluded duplicate entries, and cases with no identified *de novo* variants, retaining 13,908 trios in total. For 1,905 of these trios, variants were annotated against GRCh37. Variants for these participants were lifted over to GRCh38 using Picard (version 3.1.1)^51^. In total, we identified 979,640 variants in 13,908 individuals.

Second, we used a set of publicly available coding DNVs from exome sequencing of 31,058 rare disease trios^34^. This cohort was assembled through a collaboration between the Deciphering Developmental Disorders Study (DDD)^52^, Radbound University Medical Center (RUMC), and GeneDx. These data consist of 45,221 high confidence *de novo* calls in 23,902 individuals, annotated against GRCh37. These variants were lifted over to GRCh38 as described above, yielding 45,215 variants in 23,900 individuals.

DNVs from both cohorts were concatenated with BCFtools^45^, and annotated with allele frequencies from exome sequencing data in gnomAD v3.1.1^8^ and aggregated variant calls from whole genome sequencing of 78,195 individuals in the 100KGP. Variants were then annotated with VEP (v105)^46^. Finally, annotated DNVs were filtered on the following criteria:

1. Allele count in gnomAD v3.1.1 exomes <= 1
2. Allele frequency in gnomAD v3.1.1 exomes <= 0.0001
3. Allele count in 100KGP <= 5
4. VEP consequence annotation of “synonymous_variant”, “missense_variant”, “stop_gained”, or “frameshift_variant” in the Ensembl canonical transcript of a gene.

A small number of families are likely to have participated in both the DDD and 100KGP studies. To conservatively mitigate against “double counting” of DNVs from these individuals, we excluded from the DDD set any DNV which was also observed within the 100KGP set. 1,067 DNVs were excluded from the DDD set in this manner. We judged that participant duplication between the other cohorts was highly unlikely.

In total, 45,470 *de novo* variants (42,918 unique by chromosome, position, reference allele, and alternate allele) meeting these criteria were retained.

These variants were annotated according to the NMD region in which they fell (determined by the variant “position” in the VCF file), and with regional nonsense constraint labels. NMD regions were assigned to 46,076 DNVs. The remaining 461 DNVs are indels where the variant “position” is outside the CDS. These variants were excluded from downstream analyses. 44,174 DNVs fell in a region in which nonsense constraint could be quantified, and 29,379 DNVs fell in a region with an unambiguous nonsense constraint annotation (constrained N=15,781, unconstrained N=13,598).

### *De novo* variant enrichment

To quantify the regional enrichment of DNVs, we counted DNVs in constrained and unconstrained NMD regions. To enable direct comparisons, the absolute number of DNVs in each region was adjusted by the number of unique SNVs expected by our models in gnomAD v4.1 (above). For example, the number of unique synonymous DNVs in constrained distal regions was adjusted by the number of unique synonymous variants expected in those same regions in gnomAD. Missense variants were treated identically. Stop-gain and frameshift variants were combined, and adjusted for the number of nonsense variants expected in gnomAD. Finally, the adjusted number of variants in constrained and unconstrained regions was normalised by the adjusted number of variants in unconstrained regions. We quantified relative enrichment as the ratio of these normalised variant counts in constrained and unconstrained regions. Confidence intervals and P values for these ratios were calculated using the percentile bootstrap.

## Clinical outcome data

DNVs from the 100KGP cohort were annotated with phenotypic and clinical outcome data from the LabKey application within the Genomics England secure research environment. Specifically, these data were accessed from the “rare_disease_analysis”, “rare_disease_participant_phenotype”, and “gmc_exit_questionnaire” tables within LabKey. Diagnostic outcomes were classified as follows: a case was classed as solved where the “case_solved” value in LabKey was “yes” or “partially”; a case was classed as unsolved where the “case_solved” value in LabKey was “no”, “unknown” or absent.

## Figures

Figures 1a and 1b were created with BioRender (BioRender.com/g72m993). Transcript diagrams were created with ProteinPaint^53^. All other figures were created with Matplotlib^54^.

## Code and data availability

The analysis pipeline used to generate all results and figures is available at https://github.com/alexblakes/regional_nonsense_constraint. A separate repository for analyses undertaken in the GEL Secure Research Environment is available at https://github.com/alexblakes/gel_nmd_dnms. Regional nonsense constraint annotations and summary statistics are available for download at https://github.com/alexblakes/regional_nonsense_constraint/blob/main/data/final/regional_nonsense_constraint.tsv.

## Data Availability

Regional nonsense constraint annotations and summary statistics are available for download at https://github.com/alexblakes/regional_nonsense_constraint/blob/main/data/final/regional_nonsense_constraint.tsv

## Acknowledgements

AB is supported by a Wellcome PhD Training Fellowship for Clinicians and the 4Ward North PhD Programme for Health Professionals (223521/Z/21/Z). NW is supported by a Sir Henry Dale Fellowship jointly funded by the Wellcome Trust and the Royal Society (grant no. 220134/Z/20/Z) and a research prize from the Lister Institute. CAJ acknowledges support from MRC project grant MR/T017503/1. SB acknowledges the support of the NIHR Manchester Biomedical Research Centre. This study has been delivered through the National Institute for Health and Care Research (NIHR) Manchester Biomedical Research Centre (BRC) (NIHR203308). The views expressed are those of the author(s) and not necessarily those of Wellcome, the NIHR or the Department of Health and Social Care. This research was made possible through access to data in the National Genomic Research Library, which is managed by Genomics England Limited (a wholly owned company of the Department of Health and Social Care). The National Genomic Research Library holds data provided by patients and collected by the NHS as part of their care and data collected as part of their participation in research. The National Genomic Research Library is funded by the National Institute for Health Research and NHS England. The Wellcome Trust, Cancer Research UK and the Medical Research Council have also funded research infrastructure.

## Extended Data Table Legend

**Extended Data Table 1** Longlist of candidate disease genes which have no association with an autosomal dominant phenotype in OMIM, and carry at least 3 *de novo* nonsense or frameshift variants in a constrained NMD region in 27,762 rare disease trios.

