## Extended Data Figures for "Regional nonsense constraint offers clinical and biological insights into rare genetic disorders"

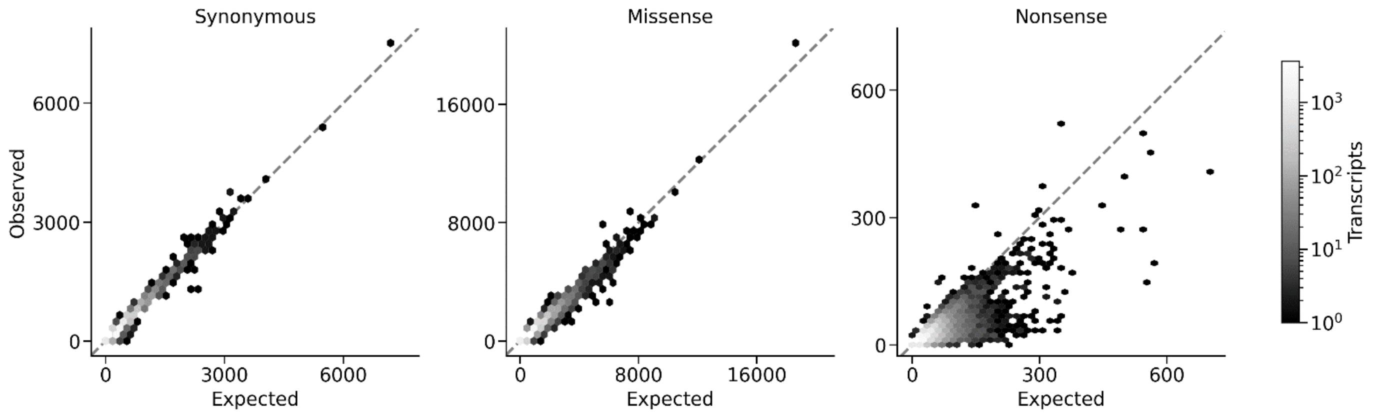


**Extended Data Figure 1** Modelling the expected number of synonymous, missense, and nonsense variants with Roulette. Hexagonal binned plots comparing the number of SNVs observed in gnomAD v4.1 versus the number expected by the null mutational model based on Roulette for 19,676 canonical protein-coding transcripts (Methods). The shading of the hexagons corresponds to the number of transcripts in each bin. The dashed grey line marks x=y. For visual clarity, *TTN* (ENST00000589042) is not plotted.

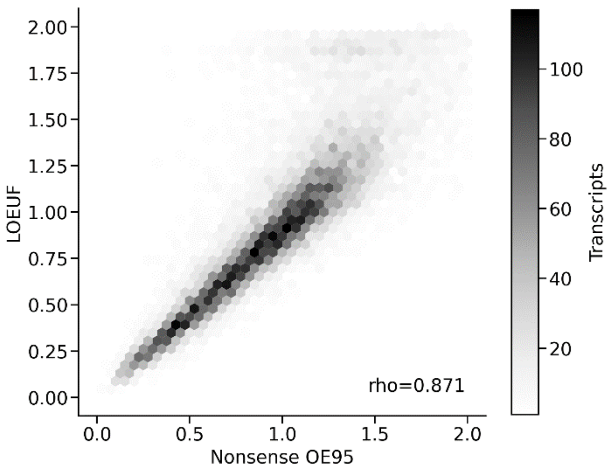


**Extended Data Figure 2** Hexagonal binned plot showing transcript-level nonsense OE95 versus LOEUF. The shading of each hexagon corresponds to the number of transcripts within that bin. Spearman's rank correlation coefficient (rho) is shown. Scores greater than 2 are not plotted for visual clarity.

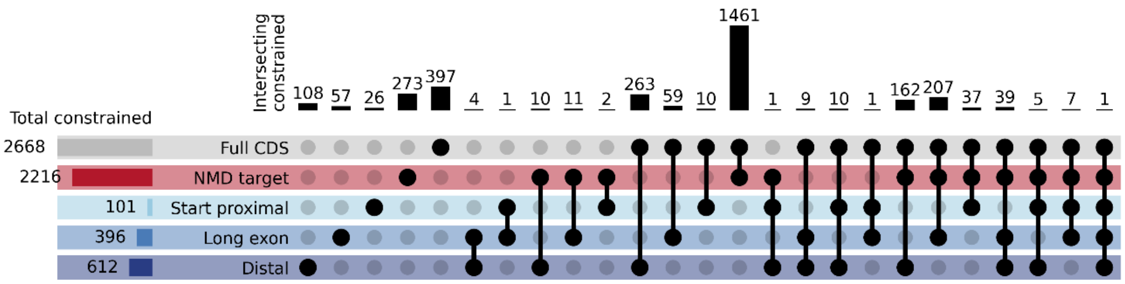


**Extended Data Figure 3** Upset plot showing the number of transcripts with regional nonsense constraint. Horizontal bars show the number of transcripts with nonsense constraint in each region. Vertical bars show the number of transcripts which harbour one or more constrained regions. Intersecting regions are shown in the matrix. For example, 273 transcripts are constrained only in NMD target regions (fourth column from left).

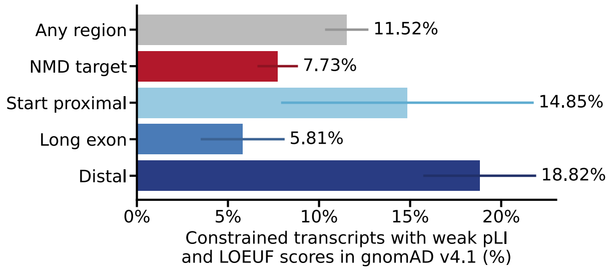


**Extended Data Figure 4** The percentage of transcripts with regional nonsense constraint which have weak pLI / LOEUF scores in gnomAD v4.1. Error bars show 95% confidence intervals.

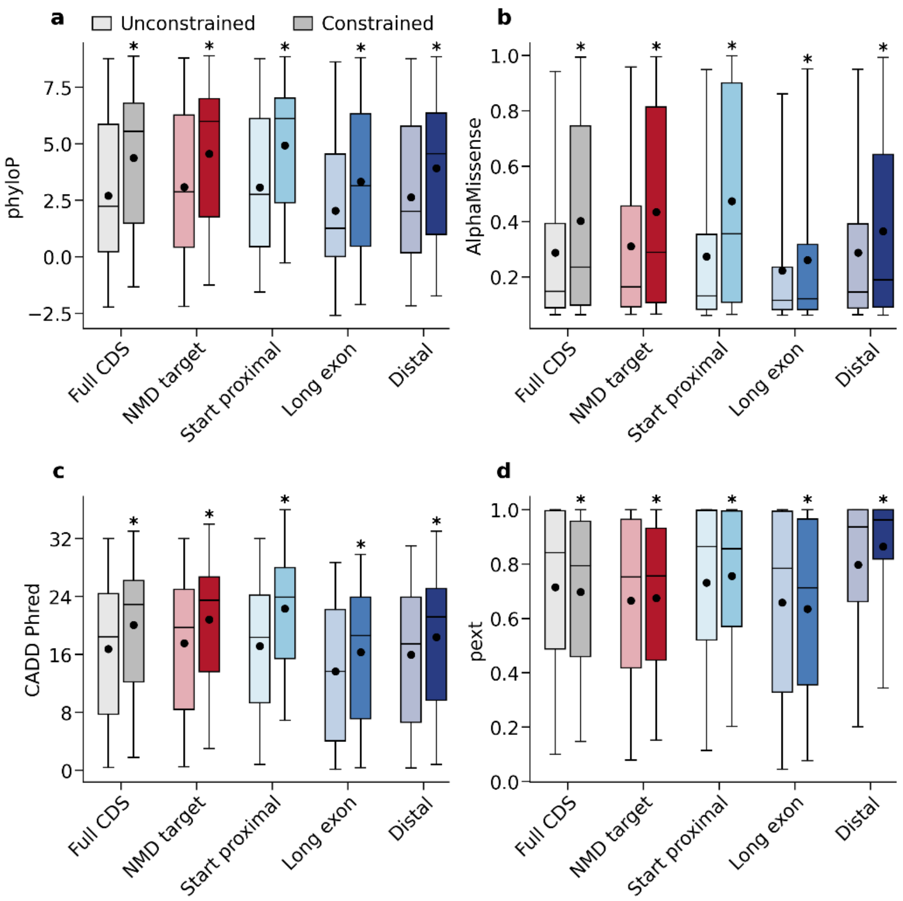


**Extended Data Figure 5** Conservation, pathogenicity prediction, and expression of sites within constrained and unconstrained regions. NMD-target regions are shown in red, NMD-escape regions in blue, and full coding sequence (CDS) in grey. Whiskers show the 5$^{th}$ and 95$^{th}$ percentiles, the box shows the 25$^{th}$ and 75$^{th}$ percentiles, and the horizontal line shows the 50$^{th}$ percentile. Closed circles show the mean. Asterisks indicate mean scores which are significantly different in constrained versus unconstrained regions (two-sided Welch's T tests, with Bonferroni correction for 20 tests at alpha=0.05, $P<10^{-8}$) **a)** Mean phyloP scores. **b)** Mean AlphaMissense scores. **c)** Mean CADD Phred scores. **d)** Mean pext scores.

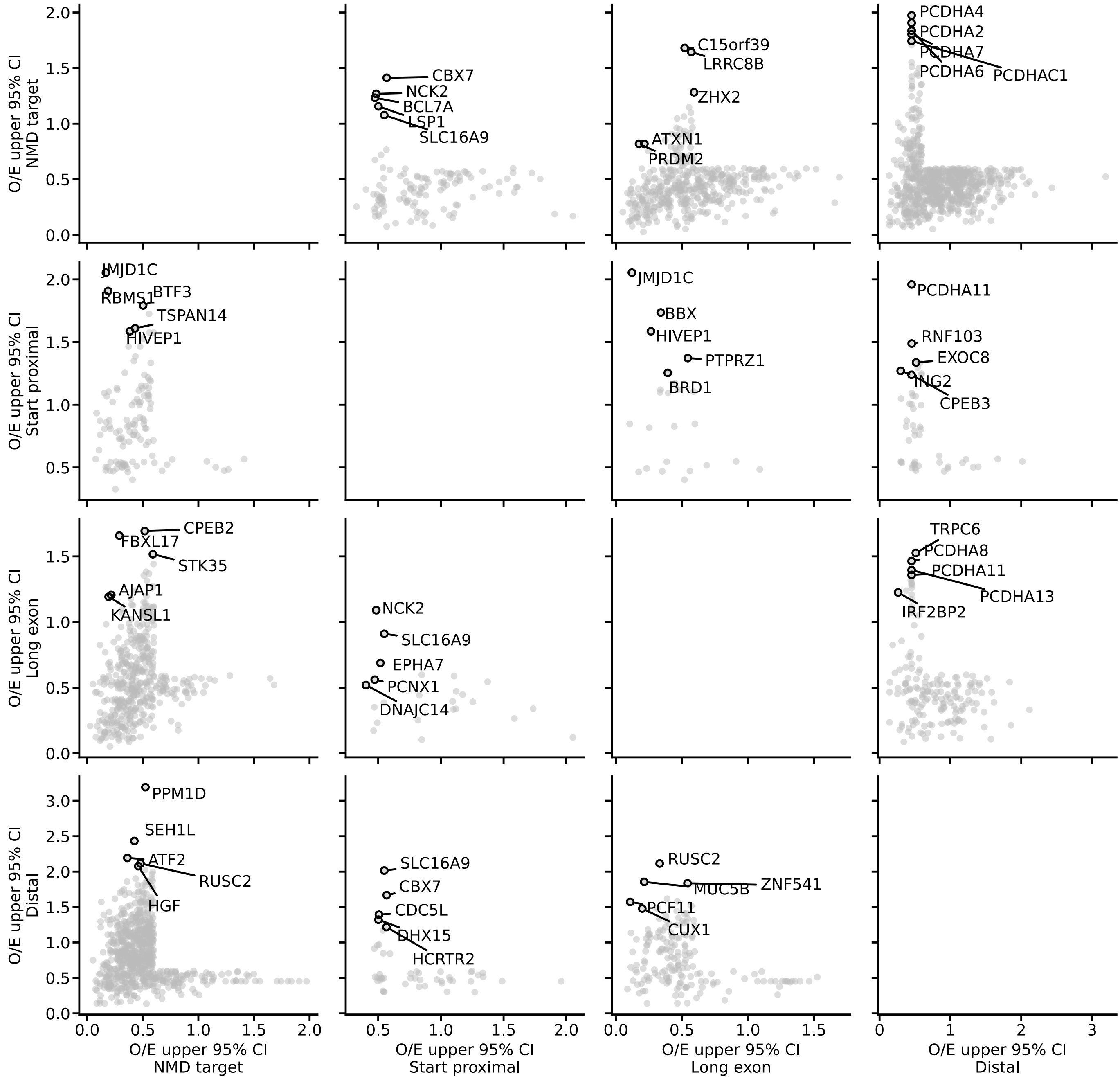


**Extended Data Figure 6** Non-uniform nonsense constraint within transcripts. Each panel compares nonsense OE95 (Methods) between NMD regions in the same transcript. Transcripts are only plotted if at least one of the regions is constrained. Within each panel, the five transcripts with the greatest absolute difference in nonsense OE95 scores are highlighted.
