## Supplementary Figures for "Regional nonsense constraint offers clinical and biological insights into rare genetic disorders"

| Region | Constraint | O/E | O/E upper 95% CI | pLI | LOEUF |
|---|---|---|---|---|---|
| **Transcript** | Constrained | 0.33 | 0.53 | 0.95 | 0.53 |
| **NMD target** | Constrained | 0 | 0.19 | | |
| **Start-proximal** | Unconstrained | 1.6 | 3.4 | | |
| **Long exon** | Unconstrained | 0.60 | 1.0 | | |
| **Distal** | NA | 0.21 | 0.90 | | |

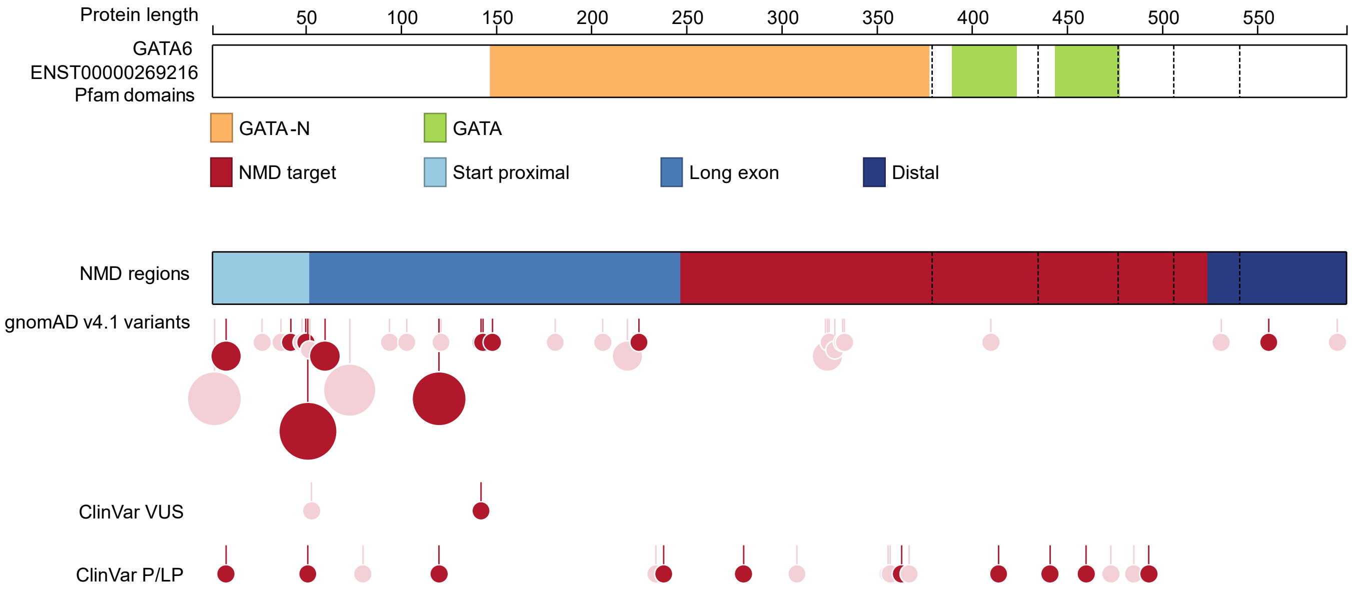


**Supplementary Figure 1** Nonsense variants are unconstrained in the start-proximal and long exon NMD-escape regions of *GATA6* (ENST00000269216), as previously described[22]. This and the other supplementary figures are arranged as in Figure 3a.

| Region | Constraint | O/E | O/E upper 95% CI | pLI | LOEUF |
|---|---|---|---|---|---|
| **Transcript** | Constrained | 0.23 | 0.35 | 1 | 0.33 |
| **NMD target** | Constrained | 0.09 | 0.19 | | |
| **Start-proximal** | Unconstrained | 1.6 | 5.4 | | |
| **Long exon** | Unconstrained | 0.73 | 1.2 | | |
| **Distal** | NA | 0.15 | 0.65 | | |

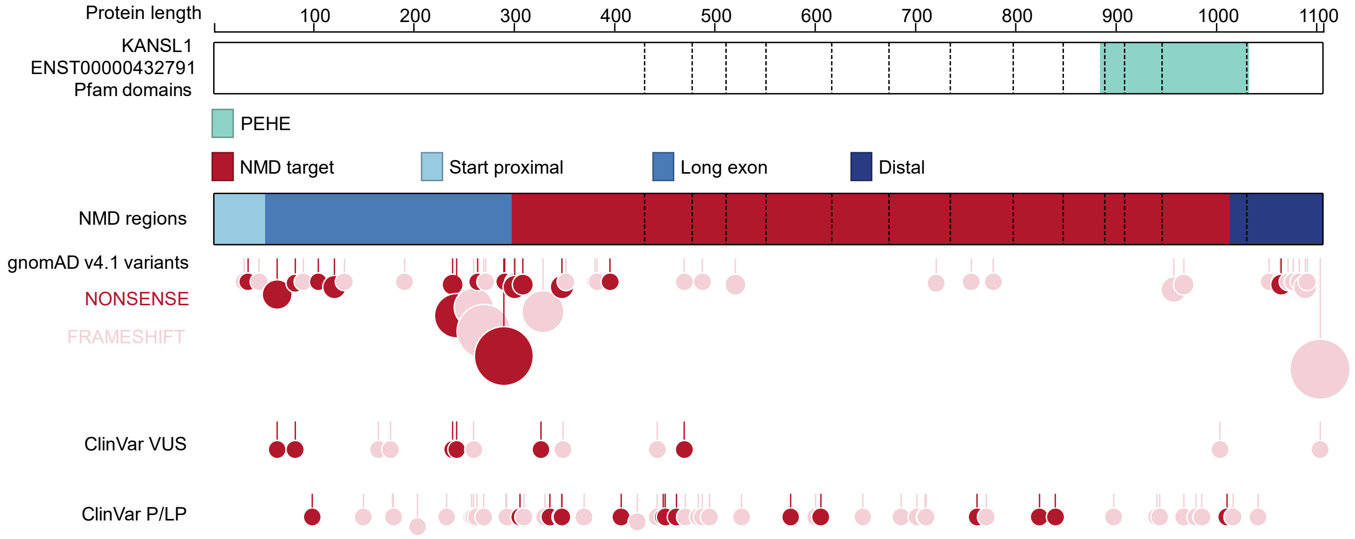


**Supplementary Figure 2** Nonsense variants appear unconstrained in the long first exon of *KANSL1* (ENST00000432791). This is likely an artefact; the 5’ CDS of KANSL1 is known to be enriched for variant annotation errors in certain haplotypes[55].

| Region | Constraint | O/E | O/E upper 95% CI | pLI | LOEUF |
|---|---|---|---|---|---|
| **Transcript** | Constrained | 0.38 | 0.56 | 0.08 | 0.68 |
| **NMD target** | Constrained | 0.13 | 0.29 | | |
| **Start-proximal** | Unconstrained | 1.7 | 3.2 | | |
| **Long exon** | Unconstrained | 1.0 | 1.7 | | |
| **Distal** | NA | 0.20 | 0.87 | | |

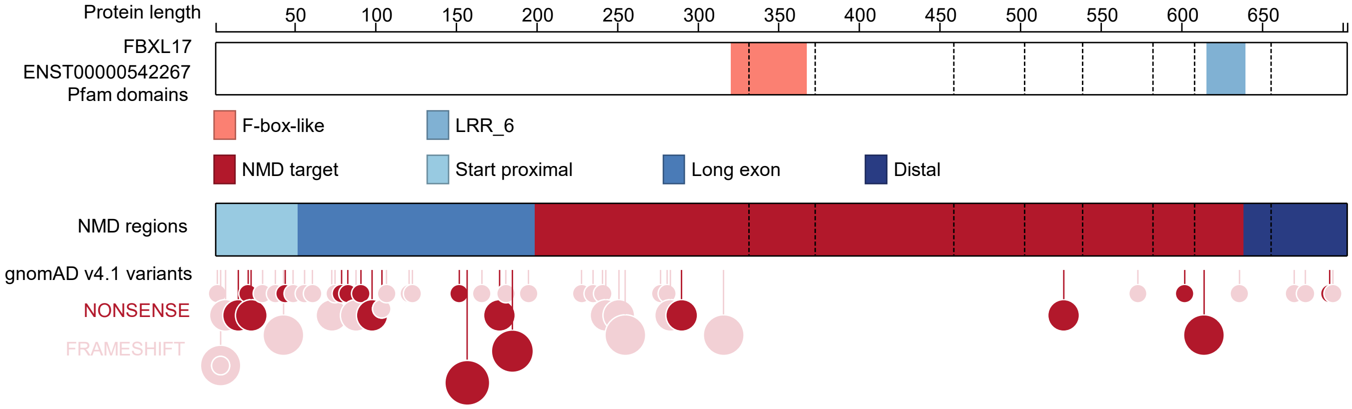


**Supplementary Figure 3** NMD-target constraint in *FBXL17* (ENST00000496714).

| Region | Constraint | O/E | O/E upper 95% CI | pLI | LOEUF |
|---|---|---|---|---|---|
| **Transcript** | NA | 0.62 | 0.94 | 0.85 | 0.64 |
| **NMD target** | Constrained | 0 | 0.34 | | |
| **Start-proximal** | NA | 0 | 1.2 | | |
| **Long exon** | Unconstrained | 0.55 | 2.0 | | |
| **Distal** | Unconstrained | 1.4 | 2.0 | | |

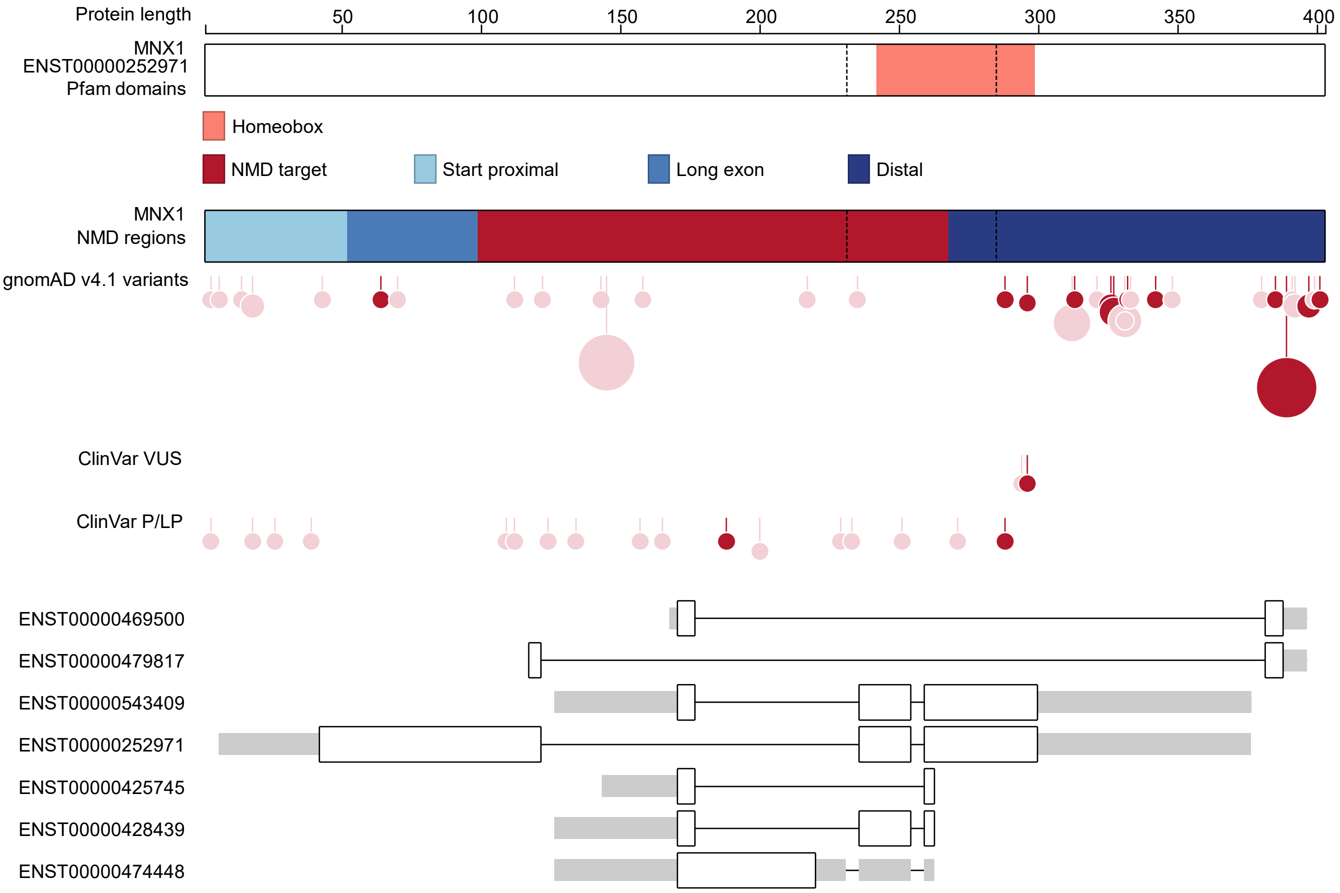


**Supplementary Figure 4** Nonsense variants are unconstrained in the distal NMD-escape region of *MNX1* (ENST00000252971).

| Region | Constraint | O/E | O/E upper 95% CI | pLI | LOEUF |
|---|---|---|---|---|---|
| **Transcript** | NA | 0.55 | 0.77 | 0.60 | 0.60 |
| **NMD target** | NA | 0.54 | 0.89 | | |
| **Start-proximal** | Unconstrained | 1.3 | 2.4 | | |
| **Long exon** | Constrained | 0.15 | 0.47 | | |
| **Distal** | Unconstrained | 0.93 | 1.6 | | |

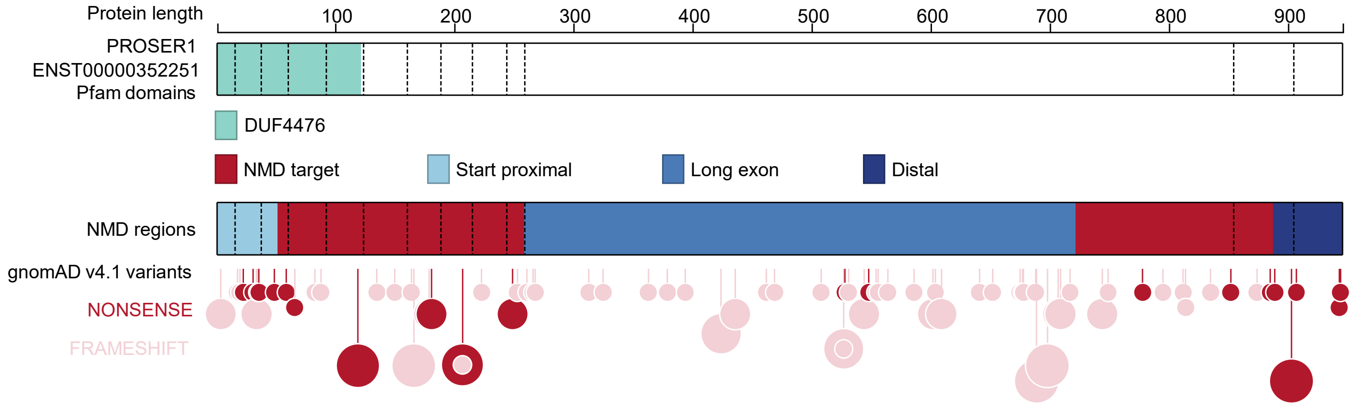


**Supplementary Figure 5** Nonsense variants are highly constrained in the long exon NMD-escape region of *PROSER1* (ENST00000352251). The bimodal distribution of nonsense variants contrasts with the more uniform distribution of frameshift variants.

| Region | Constraint | O/E | O/E upper 95% CI | pLI | LOEUF |
|---|---|---|---|---|---|
| **Transcript** | NA | 0.59 | 0.89 | 0.01 | 0.75 |
| **NMD target** | Constrained | 0.19 | 0.58 | | |
| **Start-proximal** | Unconstrained | 0.25 | 1.1 | | |
| **Long exon** | NA | NA | NA | | |
| **Distal** | Unconstrained | 1.2 | 1.8 | | |

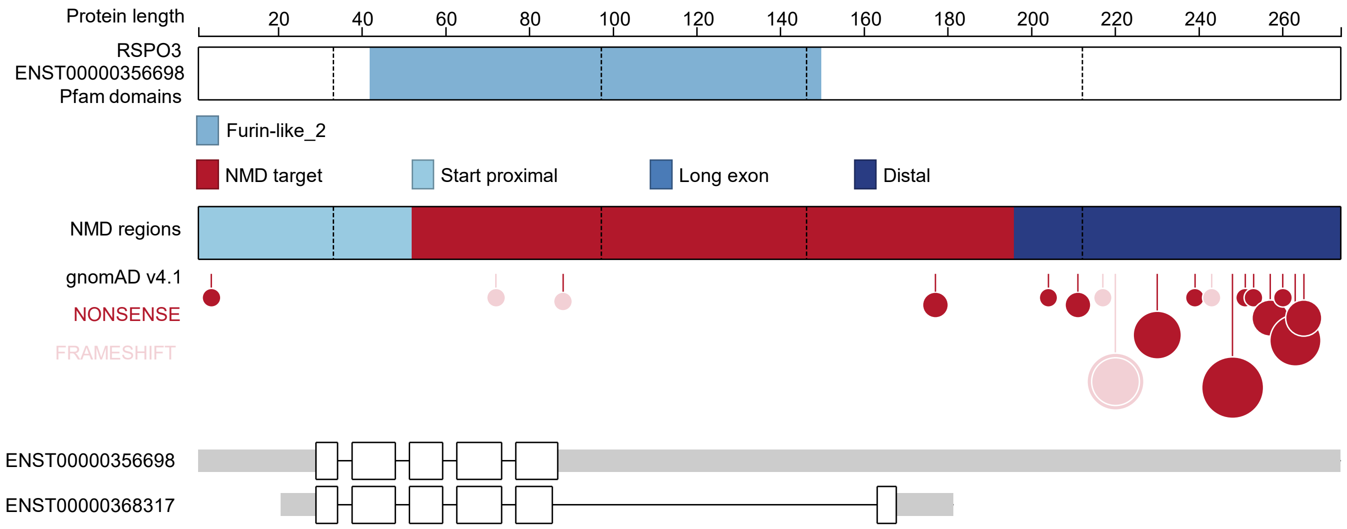


**Supplementary Figure 6** Nonsense variants are unconstrained in the distal NMD-escape region of *RSPO3* (ENST00000356698).

| Region | Constraint | O/E | O/E upper 95% CI | pLI | LOEUF |
|---|---|---|---|---|---|
| **Transcript** | NA | 0.60 | 0.96 | 0.04 | 0.77 |
| **NMD target** | Unconstrained | 0.76 | 1.4 | | |
| **Start-proximal** | Constrained | 0 | 0.57 | | |
| **Long exon** | NA | NA | NA | | |
| **Distal** | Unconstrained | 0.92 | 1.7 | | |

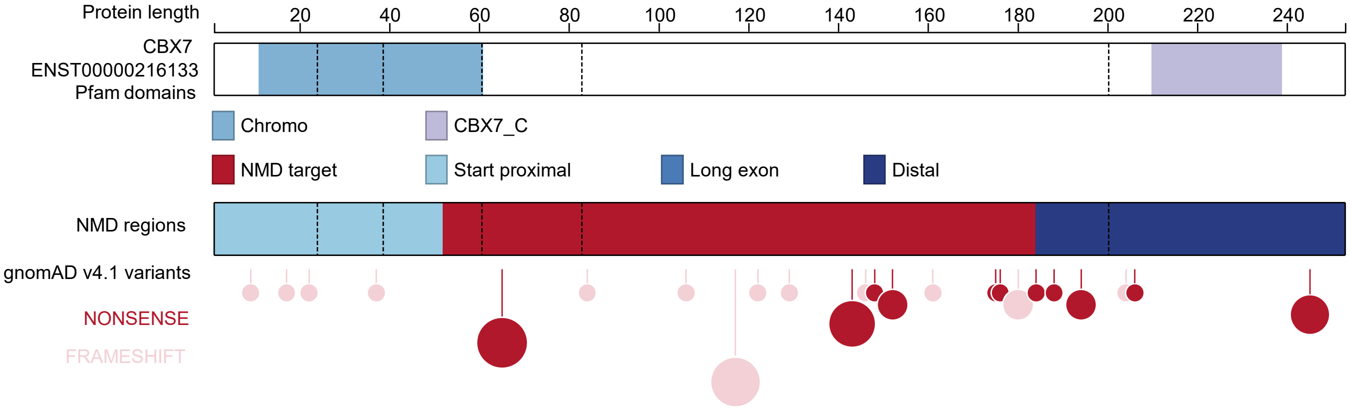


**Supplementary Figure 7** Start-proximal constraint in *CBX7* (ENST00000216133).

| Region | Constraint | O/E | O/E upper 95% CI | pLI | LOEUF |
|---|---|---|---|---|---|
| **Transcript** | Constrained | 0.29 | 0.51 | 0.41 | 0.71 |
| **NMD target** | NA | 0.19 | 0.85 | | |
| **Start-proximal** | Unconstrained | 0.66 | 1.5 | | |
| **Long exon** | Unconstrained | 0.58 | 1.2 | | |
| **Distal** | Constrained | 0 | 0.26 | | |

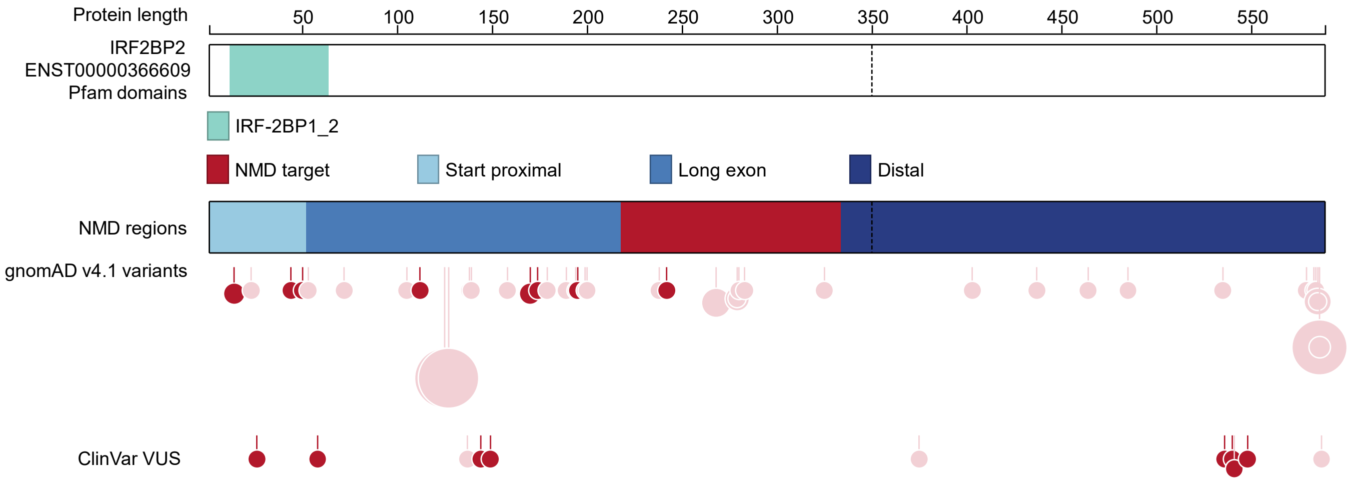


**Supplementary Figure 8** Nonsense variants are highly constrained in the distal NMD-escape region of *IRF2BP2* (ENST00000366609), but not in the 5' CDS.

| Region | Constraint | O/E | O/E upper 95% CI | pLI | LOEUF |
|---|---|---|---|---|---|
| **Transcript** | NA | 0.54 | 0.78 | 0.02 | 0.7 |
| **NMD target** | Unconstrained | 0.57 | 1.1 | | |
| **Start-proximal** | Constrained | 0 | 0.55 | | |
| **Long exon** | NA | 0.48 | 0.91 | | |
| **Distal** | Unconstrained | 1.1 | 2.0 | | |

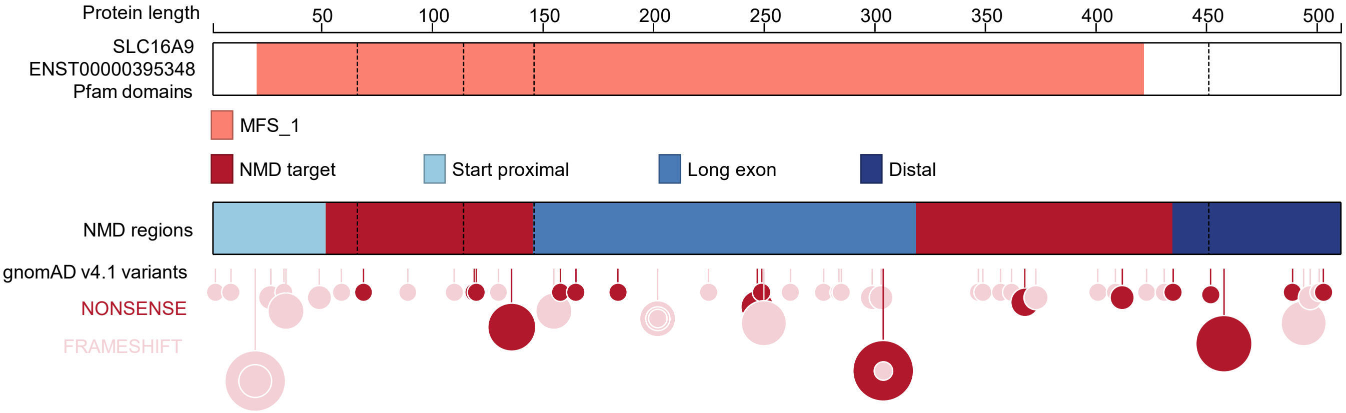


**Supplementary Figure 9** Nonsense variants are highly constrained in the start-proximal NMD-escape region of *SLC16A9* (ENST00000395348).

| Region | Constraint | O/E | O/E upper 95% CI | pLI | LOEUF |
|---|---|---|---|---|---|
| **Transcript** | NA | 0.75 | 0.97 | 0 | 0.85 |
| **NMD target** | Unconstrained | 0.57 | 1.1 | | |
| **Start-proximal** | Unconstrained | 1.4 | 2.6 | | |
| **Long exon** | Unconstrained | 0.91 | 1.2 | | |
| **Distal** | Constrained | 0 | 0.45 | | |

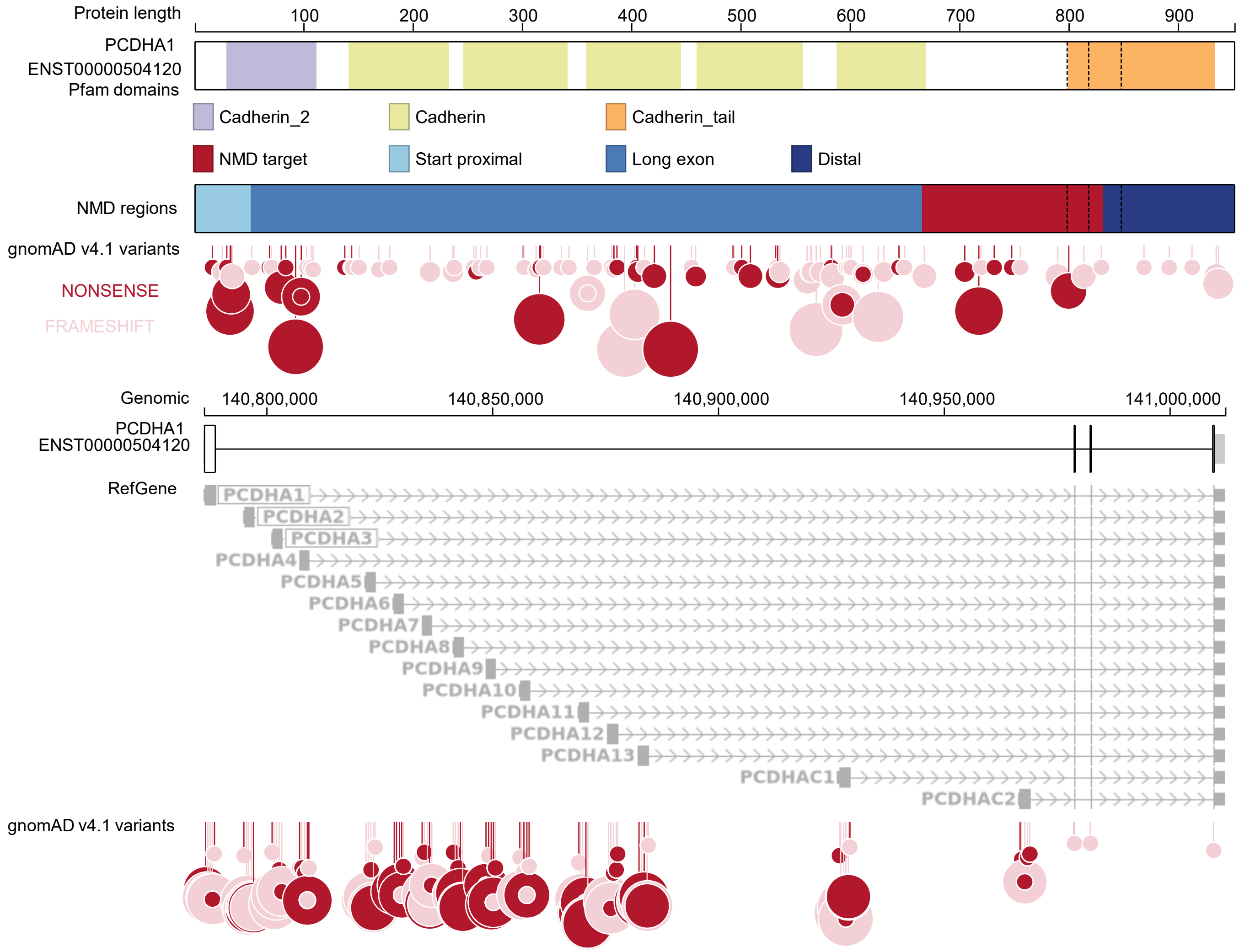


**Supplementary Figure 10** Genes in the protocadherin alpha cluster have strong nonsense constraint in the distal NMD-escape regions. The final three exons, encoding a conserved cytoplasmic domain, are shared by all genes in the cluster.

| Region | Constraint | O/E | O/E upper 95% CI | pLI | LOEUF |
|---|---|---|---|---|---|
| Transcript | NA | 0.68 | 0.90 | 0 | 0.71 |
| NMD target | Unconstrained | 0.67 | 1.2 | | |
| Start-proximal | Unconstrained | 0.58 | 2.5 | | |
| Long exon | NA | 0.72 | 1.0 | | |
| Distal | Unconstrained | 0.48 | 1.4 | | |

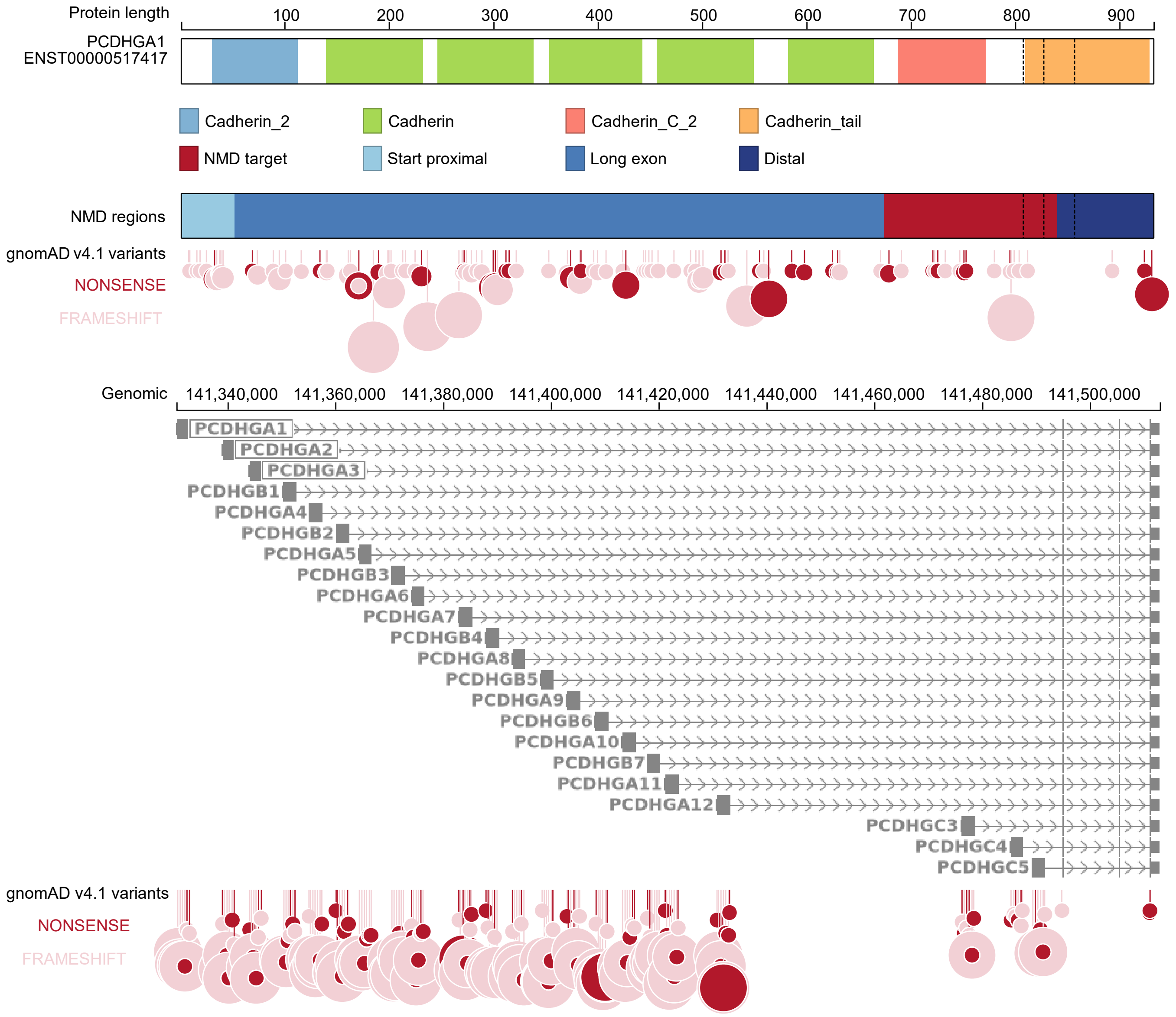


**Supplementary Figure 11** The distribution of gnomAD v4.1 nonsense variants in the protocadherin gamma cluster.
